# The Epistemonikos Database of Trials: A Comprehensive and Freely Accessible Bibliographic Source of Randomized Trials in Health

**DOI:** 10.1101/2025.11.16.25340330

**Authors:** G Rada, C Ávila-Oliver, JT Ramos-Rojas, J Vásquez-Laval, C Vergara, F Cantor-Cruz, AM Rojas-Gómez, V Veloso, C Bosio, M Bignon, S Pinto, I Silva, P Zambrano-Achig, S Fernández-Sandoval, I Jara, D Nava-Mosler, MF Verdugo-Paiva, J Peña, F Novillo, A Silva-Zapata, D Biscay

**Affiliations:** Epistemonikos Foundation, Santiago, Chile; Universidad del Desarrollo, Santiago, Chile; University of Pennsylvania, USA

**Keywords:** Randomized controlled trials, electronic databases, Epistemonikos Database of Trials (ED-Trials)

## Abstract

Randomized controlled trials (RCTs) are the cornerstone of evidence-based healthcare, providing the most reliable means of assessing the efficacy and safety of health interventions. However, their rapidly growing volume and dispersion across diverse sources, such as bibliographic databases, preprint servers, and trial registries, pose major challenges for evidence synthesis, clinical guideline development, and health technology assessment. Despite longstanding recognition of their importance and multiple initiatives to improve accessibility, there is still no comprehensive, freely available, and continuously updated resource of trial records.

To address this gap, the Epistemonikos Foundation developed the Epistemonikos Database of Trials (ED-Trials), a continuously updated repository of RCTs identified from more than 30 sources, including bibliographic databases, preprint servers, international trial registries, and other search sources. This paper describes the methods used to identify and classify trial records, outlines the key features of the search interface, and reports the database status as of October 2025.

ED-Trials now includes more than 1.5 million unique trial records, many of which are not indexed in MEDLINE or other conventional bibliographic databases. It represents a significant scholarly and technical achievement, offering a freely accessible and user-friendly resource that enables more efficient, comprehensive, and up-to-date identification of randomized controlled trials, thereby strengthening the foundation of evidence-based healthcare.

## Introduction

Randomized controlled trials (RCTs) are widely regarded as the gold standard for determining the safety and effectiveness of healthcare interventions. By randomly assigning participants to treatment groups, RCTs allow for unbiased comparisons and rigorous assessments of cause-and-effect relationships [1]. Their pivotal role in generating high-quality evidence supports clinical decision-making, systematic reviews, health technology assessments and clinical practice guidelines.

Despite their importance, comprehensively identifying all relevant RCTs and their associated data remains a significant challenge. This task requires exhaustive searches across a wide array of sources [2]. This already arduous task is exacerbated by the highly fragmented landscape of trial dissemination, which spans traditional electronic databases, clinical trial registries, preprint servers, websites, and institutional repositories [3]. This dispersion, along with the continual rise in the number of trials published annually, makes comprehensive evidence retrieval increasingly complex and resource-intensive. As such, there is an urgent need for innovative solutions to streamline access to this critical data.

Numerous initiatives have aimed to centralize RCT data to improve accessibility for research and clinical decision-making. For example, the WHO International Clinical Trials Registry Platform (ICTRP) aggregates data from multiple global trial registries [4]; the Yale Open Data Access (YODA) Project and Vivli provide platforms for sharing individual participant data submitted by researchers [5],[6]. The Cochrane Central Register of Controlled Trials (CENTRAL), one of the most comprehensive efforts, compiles RCTs from a range of sources, including bibliographic databases and trial registries [7]. However, these initiatives face significant limitations: they cover a restricted set of sources, rely heavily on manual work, and some impose access restrictions or fees.

In response to these challenges, the Epistemonikos Foundation has developed the **Epistemonikos Database of Trials (ED-Trials)** (https://trials.epistemonikos.org/), a free, comprehensive repository of RCTs in health. This database builds on the proven strategies used in the Epistemonikos Database of Systematic Reviews [8] and the COVID-19 L.OVE Platform [9]. Its overarching goal is to consolidate all relevant RCTs in health and their associated data into a single, freely accessible resource. By reducing reliance on time-consuming and fragmented searches, ED-Trials aims to facilitate more efficient access to essential evidence.

This paper describes the methods used to identify trial records in ED-Trials, presents the key features of the search interface, and reports the results as of October 2025, underscoring its potential as a global public good for evidence-based healthcare.

## Methods

We built our methods by following or adapting the Preferred Reporting Items for Systematic Reviews and Meta-Analyses (PRISMA) [10].

### Inclusion criteria

We defined a randomized trial as an experimental study design in which participants or other units of randomization are prospectively assigned, using random allocation, to two or more alternatives or to different sequences of interventions, to compare their effects [1].

We include any document related with the design, planning, conduct, or results of randomized trials. This includes trial registry entries, published protocols, preliminary results, final outcomes, post-hoc analyses, and interim analyses of specific randomized trials, regardless of trial phase, sample size, population studied, intervention type, comparator, or outcome measures.

The operational criteria for inclusion in the ED-Trials are:

- Randomized allocation: The study must explicitly report random allocation.
- Controlled design: The trial must compare at least one alternative against a control (e.g., placebo, no intervention, standard treatment) or another active alternative.

No restrictions are applied regarding the topic or field of the trial. Trials are eligible irrespective of their completion or publication status, including those reported in conference proceedings or grey literature. No limits are placed on language or publication date.

Studies are excluded if they:

- Are explicitly identified as quasi-experimental, observational, or other non-randomized designs.
- Report methods or results of non-randomized study designs conducted before, during, or after an RCT, where the results are not attributable to the randomized groups.

### Search and retrieval

#### Electronic sources

ED-Trials is maintained through systematic searches of multiple electronic sources. Most searches are conducted on a daily basis through automated processes, using publicly available APIs, RSS feed subscriptions, or .csv file parsing. Paywalled sources are manually searched on a weekly schedule, and when a unique article is identified (i.e., not available through any public source), only the bibliographic citation is added to ED-Trials.

As of October 2025, the database routinely harvests content from more than 30 electronic databases, preprint servers, and trial registries using the following strategy, adapted to the syntaxis of each database: randomi* OR RCT OR placebo* OR trial OR “controlled-trial” OR randomly*. A comprehensive list of sources is provided in **Appendix 1**. A continuously updated version of this list is available at: https://trials.epistemonikos.org/en/methods.

#### Other search sources

To identify RCTs that may not be captured through electronic sources, the following additional approaches are followed:

- Utilizing the *linker* tool from the Epistemonikos Database of Systematic Reviews (operational since 2009) which maps systematic reviews to their included studies [8].
- Incorporating references submitted by clinical experts or users through email, contact forms, or social media channels.

#### Search strategy

A comprehensive search strategy was developed and adapted for the specific syntax of each electronic source (see **Appendix 1** for detailed search strategies in each source). For example, the following search string is run in PubMed: “Clinical Trial” [Publication Type] OR randomi* OR RCT OR placebo* OR trial OR “controlled-trial” OR randomly*.

#### Deduplication and quality assurance

References retrieved from electronic sources are automatically deduplicated using a proprietary algorithm that compares both unique identifiers (e.g., database IDs, DOIs, trial registration numbers) and bibliographic citation data (including author names, journal title, publication year, volume, issue, page numbers, article title, and abstract). This automated process is supplemented by a manual review performed by the Epistemonikos Foundation’s quality assurance team. Records missing essential bibliographic elements (such as author, title, source, year, or abstract), containing errors (e.g., HTML characters), or lacking an English version of the title and abstract are automatically flagged and subsequently corrected through automated or manual procedures by the quality assurance team.

### Study selection

#### Automated methods

Multiple automated methods are used to identify and select studies for inclusion in ED-Trials.

Automated Method 1: The initial classification process relies on a highly sensitive deep neural network classifier, the development and validation of which have been previously described [11].

Automated Method 2: To enhance specificity, a second classifier is applied to the articles selected by Method 1 and, if available, to those tagged as RCTs in the original source. This method uses a large language model (LLM) prompt, developed through iterative refinement and evaluated across various models. The final prompt, detailed in the **Appendix** 2, has been implemented using the Gemini 2.0 Flash model as of October 2025.

Automated Method 3: For sources that expose structured metadata through APIs, we extract information directly when the source explicitly tags studies as RCTs.

Automated methods 1 and 2 are applied only to records that include an abstract.

#### Semi-*automated methods*

The incorporation of trials from systematic reviews in the Epistemonikos Database of Systematic Reviews relies on a combination of automated and semi-automated methods [8]. First, systematic reviews that potentially include RCTs are identified using the same process described above for RCT detection. Each review is then checked manually to determine whether it includes only RCTs or a mix of RCTs and non-RCTs. When a review includes only RCTs, all its studies are automatically tagged as such; when both types are present, human extraction is performed.

#### Human validation

Targeted manual validation is carried out by the Epistemonikos methods team in collaboration with a network of trained reviewers. This process focuses on specific groups of articles, including:

- Records retrieved through Boolean searches that lack an abstract and were not captured by any automated method.
- Records showing discrepancies between automated and human classifications.
- Records with inconsistencies between classifications assigned in systematic reviews and those generated by automated methods or human reviewers.
- Records with disagreements between different human reviewers.

### Database features

ED-Trials is designed to support users, particularly those involved in evidence synthesis, by integrating a suite of advanced features that promote efficient and accurate trial retrieval and management. These features aim to streamline the identification of relevant randomized controlled trials and facilitate their integration into systematic reviews, guidelines, and other forms of evidence synthesis.

#### Advanced search interface

The platform includes a highly functional and intuitive advanced search interface **(see Figure 1)**, built to support both novice and expert users. It enables single-line and multi-line search modes, allowing users to construct queries with precision and flexibility. The interface supports Boolean logic and field-specific queries.

**Figure 1.**
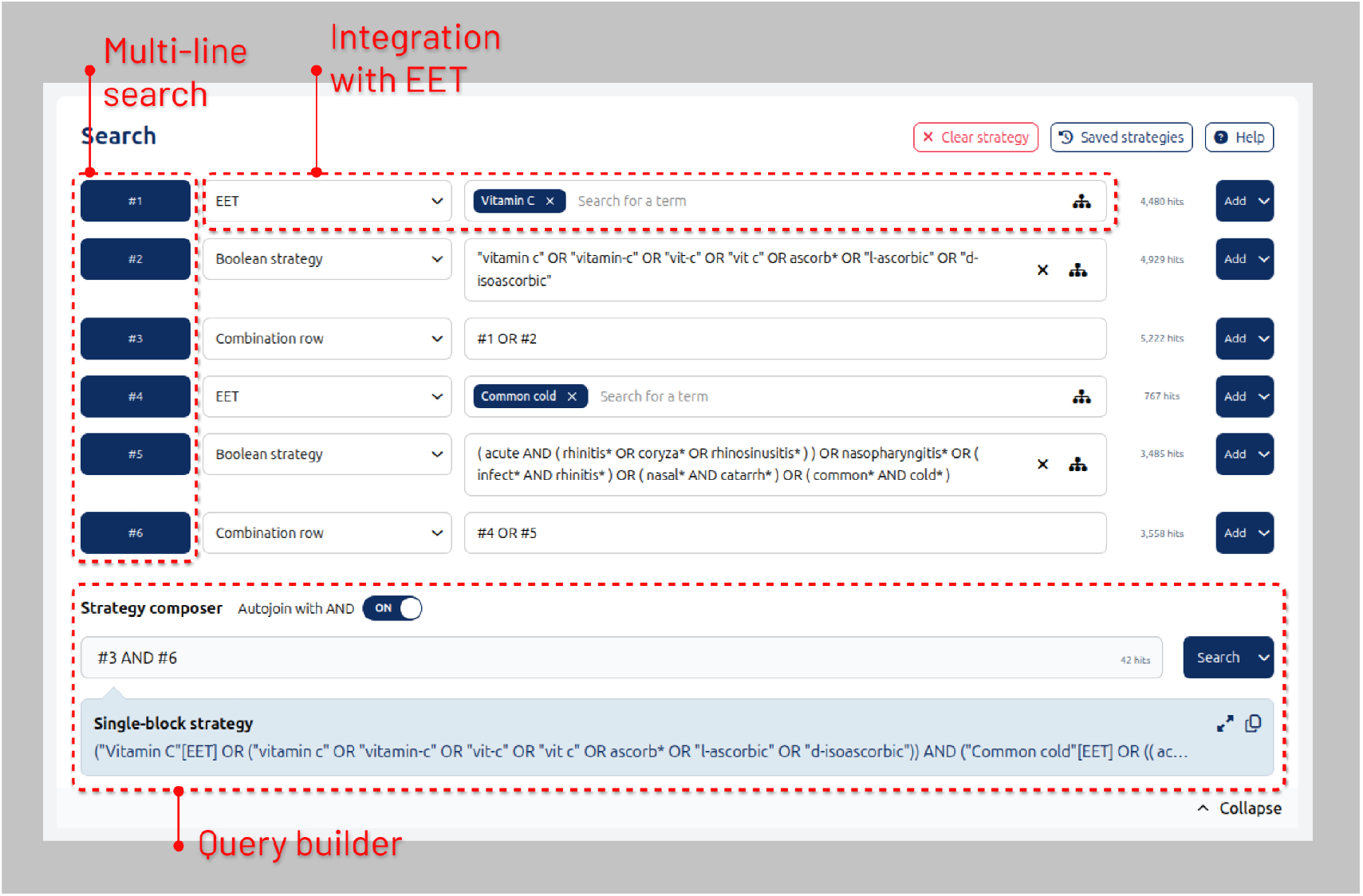
ED-Trials advanced search interface

Central to the advanced search functionality is the integration of the Epistemonikos Evidence Taxonomy (EET), developed to support consistent and precise classification of health-related concepts. The EET allows users to compose semantically rich queries that improve both sensitivity and specificity in retrieving relevant studies [12].

#### Automated Boolean strategy creator

To further support users, particularly those conducting systematic reviews and other structured evidence syntheses, the database offers an Automated Boolean Composer (**See Figure 2)**. This innovative tool automatically inserts high-quality, pre-formulated search strategies directly into the search interface. These strategies are built on tested Boolean logic and use standardized terms from the EET to ensure comprehensive coverage of a given concept or condition [12].

**Figure 2.**
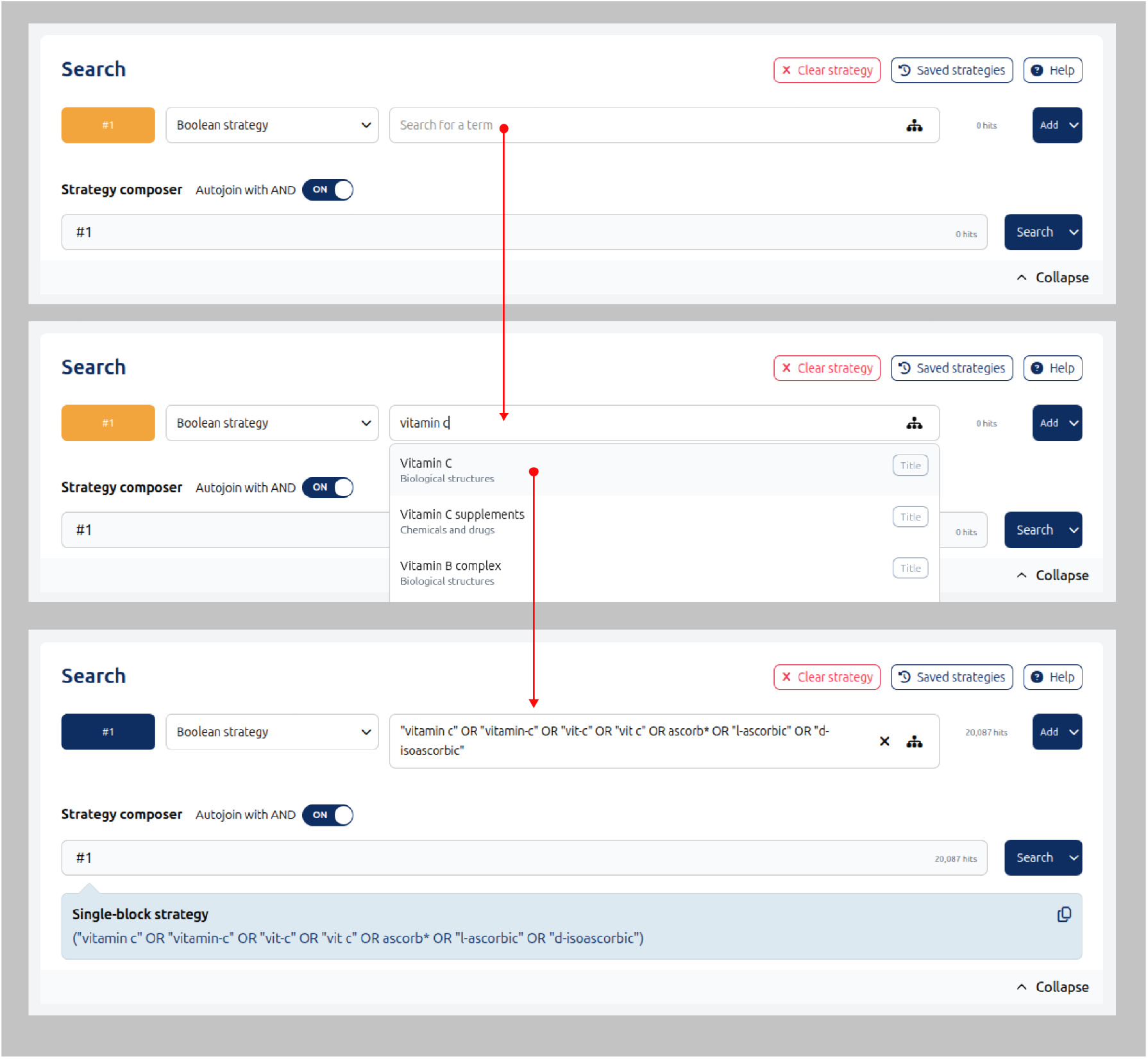
Automated Boolean Composer

The generated strategies serve as editable drafts that can be refined or customized based on the specific focus of a review or project. This feature is particularly valuable for users without advanced search skills, as it reduces the potential for common errors and enhances reproducibility.

## Results

As of October 2025, more than 7.7 million records have been retrieved from the different sources. After removing duplicates and applying the selection process, the total number of included randomized trials in ED-Trials is 1,501,729. A flow diagram summarizing the study identification process is presented in **figure 3**.

**Figure 3.**
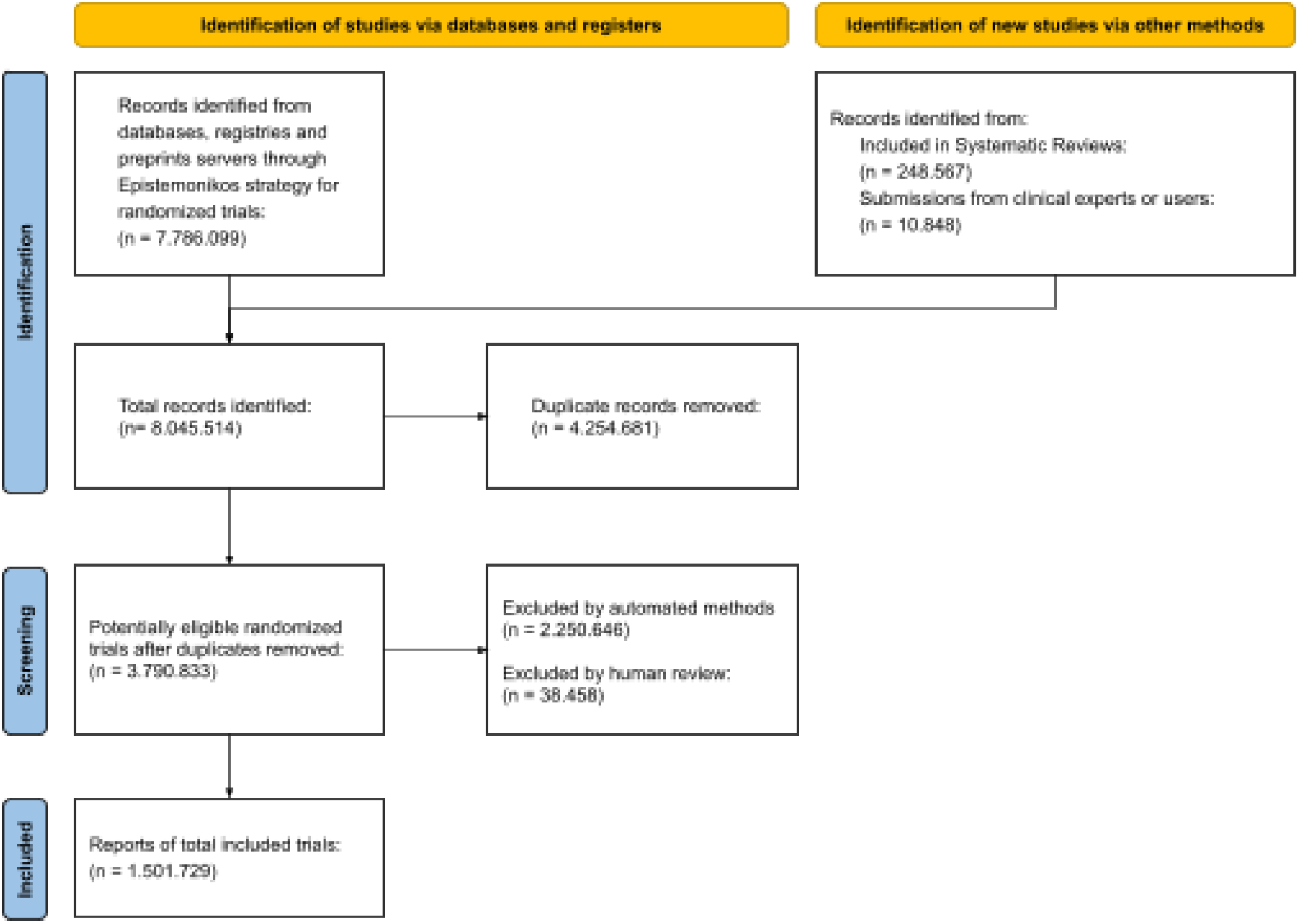
PRISMA flow diagram of the study identification process

The number of randomized trials published globally since 1975 has experienced a marked and sustained expansion since 1975, with a pronounced acceleration beginning in the early 2000s, particularly driven by increases in journal publications and trial registry records (**Figure 4**). The total volume of RCTs continued to rise steadily through the 2010s, reaching peak levels in the early 2020s, although a decline is observed in 2024–2025 (with 2025 still incomplete). Registry records, which were almost nonexistent before 2000, show a major rise beginning around 2004, reflecting both the emergence of trial registration norms and increased transparency requirements. Other publication types remain comparatively small but show gradual growth.

**Figure 4.**
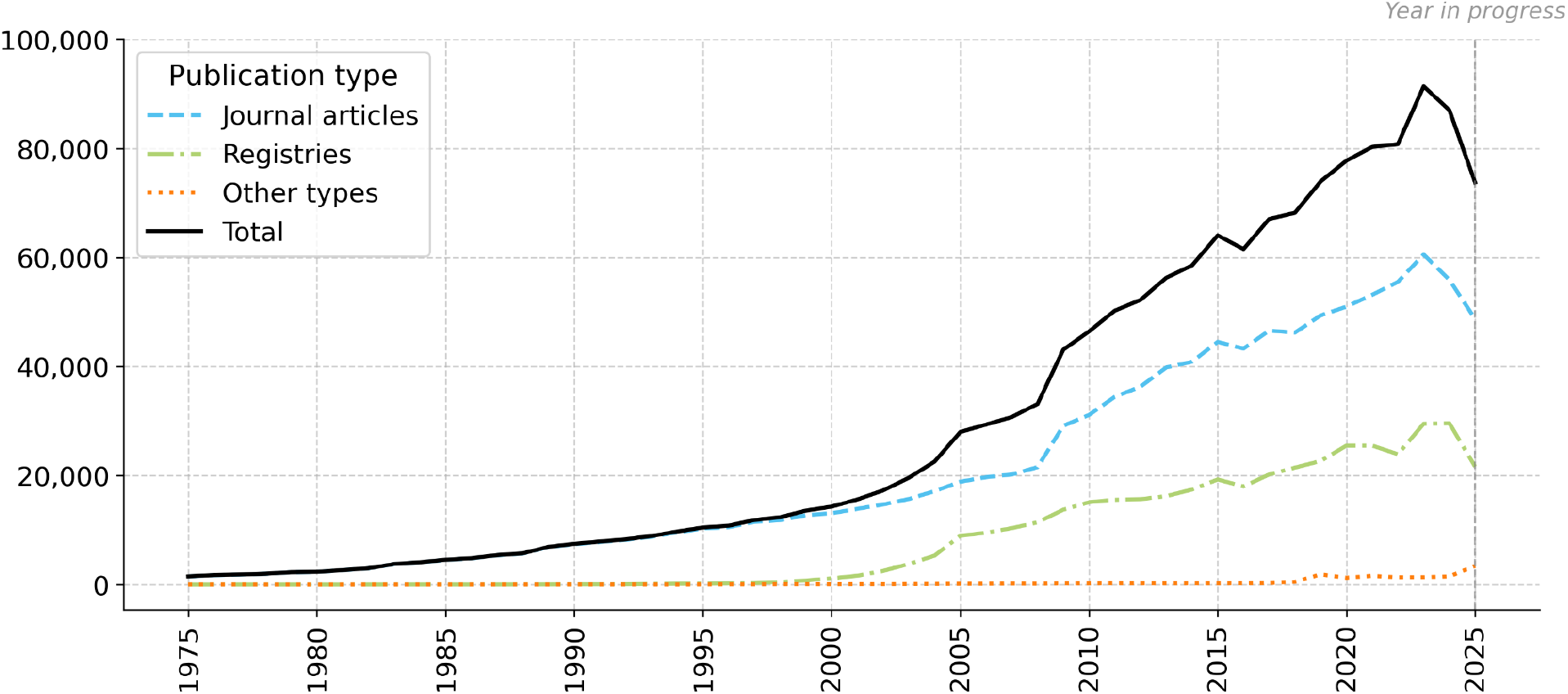
Number of randomized trials per year

Over the past five years, an average of 235 randomized trial records have been published each day. Of these, 71.7% correspond to journal articles, 27.2% to trial registry entries, and 1.1% to other publication formats such as preprints or non-journal reports **(Figure 5)**.

**Figure 5.**
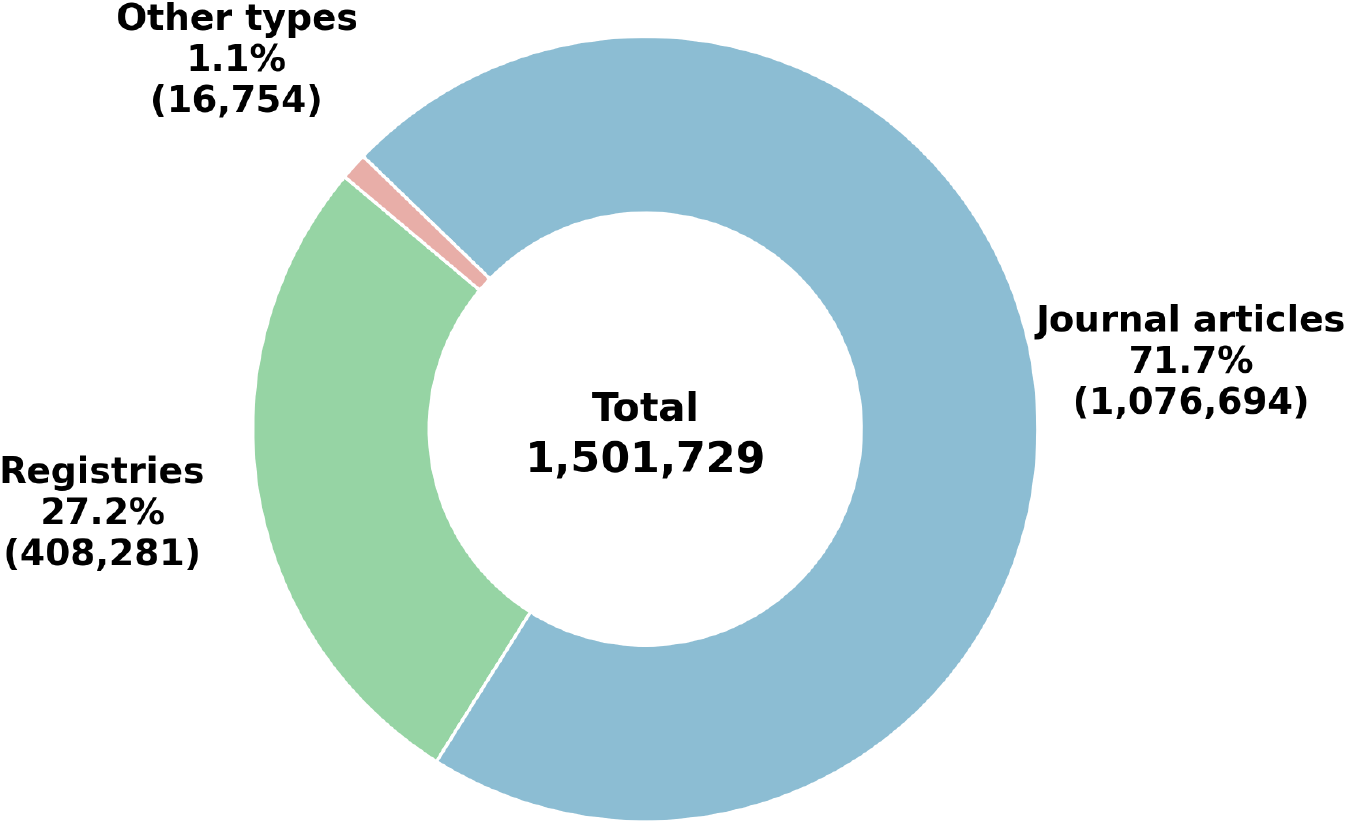
Records included in ED-Trials by publication type

**Figure 6.**
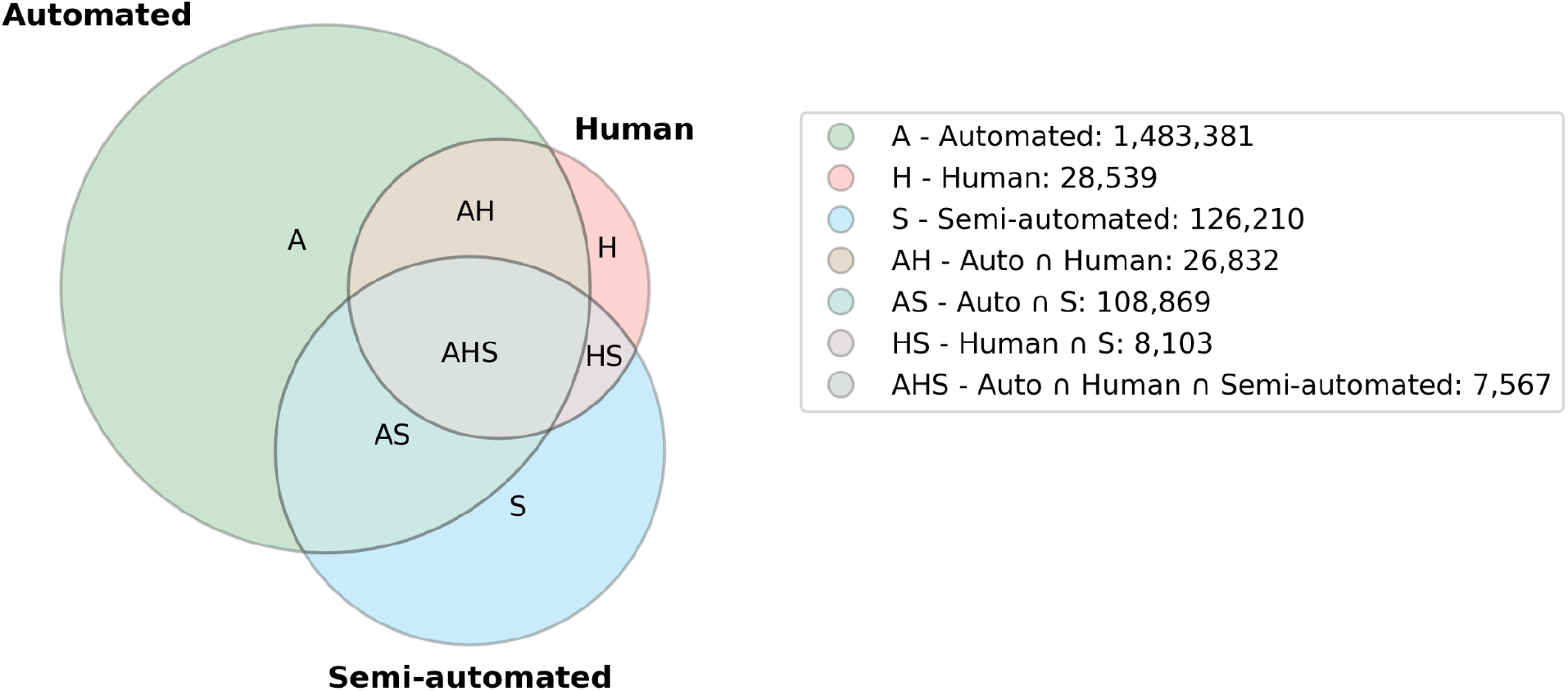
Records included in ED-Trials by classification method

In terms of the method used to classify the articles, 1,483,381 articles have been selected using automated methods, 126,210 through semi-automated approaches, and 28,539 through human classification. Although there is substantial overlap across all three classification strategies, 1,355,247 trials were identified exclusively by automated methods, 16,805 exclusively by semi-automated methods, and 1,171 exclusively by human review.

## Discussion

The Epistemonikos Database of Trials represents a major advancement in the accessibility to randomized controlled trial evidence. Through systematic and continuous searches across an unprecedented range of electronic databases, preprint servers, and international trial registries, supplemented by the inclusion of trials identified through systematic reviews and other sources, ED-Trials addresses the long-standing issue of fragmented and dispersed trial data.

This comprehensive and centralized repository of RCTs significantly enhances the capacity for evidence synthesis and informed decision-making in health care. In doing so, it fulfills a vision long advocated by pioneers of evidence-based medicine, such as Archie Cochrane [13]. Building on this foundation, the database’s free-access model further reinforces its role as a global public good. By facilitating access to high-quality evidence, ED-Trials empowers researchers, policymakers, clinicians, and other stakeholders across diverse settings to engage in evidence-informed healthcare decisions, thereby contributing to more equitable and effective health systems worldwide.

The novelty of ED-Trials lies not only in its extensive coverage but also in its suite of innovative features designed to optimize evidence retrieval and use. The advanced search interface, enhanced by the Epistemonikos Evidence Taxonomy and an automated Boolean strategy generator, improves the rigour and efficiency of search strategies. In addition, its direct integration with evidence synthesis platforms allows researchers, such as systematic review authors, or guideline developers, to seamlessly transition from study identification to analysis. This streamlined workflow is particularly valuable for supporting living evidence processes and other dynamic approaches to evidence generation and use.

Several limitations must be acknowledged. First, although ED-Trials is designed to be a comprehensive repository of randomized controlled trials, its actual sensitivity has not yet been fully established. Formal validation studies are currently underway to assess its completeness and accuracy in identifying RCTs across different fields. These evaluations are critical to determine whether ED-Trials can reliably replace the labor-intensive practice of searching across multiple databases, a resource-intensive step in evidence synthesis.

Second, a technical limitation is the potential inclusion of studies incorrectly identified as randomized trials (i.e., false positives). Despite the use of state-of-the-art classification technologies and a rigorous human validation process, misclassification remains a possibility.

Lastly, a major limitation concerns long-term sustainability. The Epistemonikos Foundation, while having demonstrated a strong track record in maintaining major evidence resources, such one of the world’s largest databases of systematic reviews since 2012 [14] and the most comprehensive COVID-19 evidence database since 2020 [9], is a self-funded organization. The continued maintenance and development of ED-Trials will require sustained resources and support, which are not yet guaranteed.

## Conclusion

ED-Trials is a novel, comprehensive, and freely accessible database of randomized controlled trials. Through its systematic search approach, integration of diverse sources, and advanced search and connectivity features, it aims to eliminate the need for laborious multi-source searching. The ED-Trial serves as a critical resource for systematic review producers, guideline developers, and researchers in general, significantly enhancing the efficiency of evidence identification and accelerating the production of high-quality evidence syntheses, thereby contributing to evidence-informed decision-making in healthcare globally.

## Data Availability

All data produced in the present study are available upon reasonable request to the authors

https://trials.epistemonikos.org/

## Data Availability

All data produced in the present study are available upon reasonable request to the authors

https://trials.epistemonikos.org/

## Acknowledgements

We thank Jaime Cerda and Jorge Dagnino who suggested the name Epistemonikos after long and stimulating conversations. This project would not have been possible without the support of all the organisations that have provided funding, a network of volunteers who have contributed with invaluable work.

## Contributions

Gabriel Rada: Conceptualization, Writing - Original Draft preparation, Validation. Camila Ávila-Oliver: Writing - Review & Editing, Validation. José Tomás Ramos-Rojas: Writing - Review & Editing, Validation. Juan Vásquez-Laval: Writing - Review & Editing, Software. Camilo Vergara: Writing - Review & Editing, Software. Francy Cantor-Cruz: Data Curation. Ana María Rojas-Gómez: Data Curation. Valentina Veloso: Data Curation. Claudia Bosio: Data Curation. Magdalena Bignon: Data Curation. Sebastián Pinto: Data Curation. Iván Silva: Data Curation. Paula Zambrano-Achig: Data Curation. Sergio Fernández-Sandoval: Data Curation. Iván Jara: Software. Daniel Nava-Mosler: Software. María Francisca Verdugo-Paiva: Validation, Data Curation, Javiera Peña: Data Curation. Francisco Novillo: Data Curation. Álex Silva-Zapata: Software. Diana Biscay: Writing - Review & Editing, Validation.

## Authors’ information

Epistemonikos (from the Greek “what is worth knowing”) is an independent, non-profit organization dedicated to providing reliable evidence for healthcare decision-making. One of its core areas of work is the development of systems that accelerate the identification, processing, and visualization of scientific evidence.

## Appendix 1. Search sources

### Electronic databases

- MEDLINE/PubMed
- EMBASE
- CINAHL
- LILACS
- IRIS WHO
- PsycINFO
- Scopus

#### Preprint servers

- medRxiv/bioRxiv
- SSRN
- ResearchSquare

#### Trial Registries

- ClinicalTrials.gov
- ISRCTN registry
- EU Clinical Trials Register
- Iranian Registry of Clinical Trials
- Australian New Zealand Clinical Trials Registry
- International Clinical Trials Registry Platform
- Chinese Clinical Trial Register
- Brazilian Registry of Clinical Trials
- Pan African Clinical Trials Registry
- Lebanese Clinical Trials Registry
- Registro Público Cubano de Ensayos Clínicos
- Overview of Medical Research in the Netherlands
- Clinical Trials Peruvian Registry
- Korean Clinical Trials Database
- UMIN Clinical Trials Registry
- Japan Primary Registry Network
- Japan Pharmaceutical Information Center
- German Clinical Trials Register
- Clinical Trials Registry - India
- Sri Lanka Clinical Trials Registry
- Thai Clinical Trials Registry
- International Traditional Medicine Clinical Trial Registry
- Clinical Trials Information System

## Appendix 2. LLM prompt for randomized controlled trials

### System prompt

~~~
## ROLE: You are an expert health scientist who has been asked to review the article.
Keep in mind that the response will be read by an automated script so the answer must be in JSON
format as shown in the provided example
Use this context to classify the article:
## CONTEXT:
### TITLE: []
### ABSTRACT: []
## EXAMPLES:
{‘responses’: [{‘id’: ‘<id>‘, ‘value’: ‘<yes, no or unclear>‘}, {‘id’: ‘<id2>‘, ‘value’: ‘<yes, no or unclear>‘}]}
~~~

#### User prompt

~~~
## TASK: Identify with YES/NO/UNCLEAR if the article has any of the following sub-classifications. It’s required that you answer all the questions. If you are not sure, please answer UNCLEAR. ONLY answer YES if the article is a clear example of the sub-classification.
## QUESTIONS:
[
  {
   “id”: “5f15c47469c00e5e2d7fdc25”,
     “name”: “randomized trial”,
       “inclusion_criteria”: “Retrieve scientific articles that assess the design, planning, conduct, or results of randomized trials conducted in human subjects, or in animals/non-human subjects. This includes trial registry entries, published protocols, preliminary results, final outcomes, post-hoc analyses, and interim analyses of specific randomized trials, regardless of trial phase, sample size, population studied (human or relevant non-human), intervention type, comparator, or outcome measures. Exclude systematic reviews, meta-analyses, narrative reviews, scoping reviews, and other review articles that synthesize or analyze data from multiple randomized trials. Prioritize sensitivity to minimize the risk of excluding potentially relevant primary literature that directly reports on a specific randomized trial’s planning or findings.”
   }
  ]
## RESPONSE:
~~~

